# Thermal Effect On The Persistence Of SARS-CoV2 Egyptian Isolates As Measured By Quantitative RT-PCR

**DOI:** 10.1101/2020.10.13.20211771

**Authors:** M.G. Seadawy, A.F. Gad, M.F. Elhoseny, B. ELharty, M.S. EL Desoky, Y.A. Soliman

## Abstract

Coronavirus pandemic that caused by severe acute respiratory syndrome Coronavirus 2 (SARS-CoV-2) appeared in China in 2019 then spread all over the world .COVID-19 firstly appeared in Egypt in Feb 2020. Studies on the thermal stability of the virus is crucial proper specimens’ transportation for molecular study. Oropharyngeal swabs were taken from recently infected military people with COVID-19 from Egypt during April 2020. Samples were aliquoted and the thermal stability of the virus was measured using quantitative real Time RT-PCR for samples treated at different temperature ranges from 20 °C to 70 °C for 2,4and 6 hours. Results shown that inactivation of the virus and significant reduction in the ΔCq values begin at 40 °C/4h. Complete virus inactivation and loss of ΔCq values were seen at 50 °C/6h and 60 °C. Tested samples showed no significant difference in thermal stability at any temp/time combinations tested.

## Introduction

A novel coronavirus (SARS-CoV-2) also known as COVID-19 has recently emerged and first recorded in Wuhan city, China in December 2019 (*Ali, et al*., *2019*) with more than 5.7 million infected patients and deaths toll exceeds 357K by the end of May 2020 (*WHO, 2020 report -130*).

The first case reported of COVID-19 pandemic in Egypt confirmed on 14 Feb 2020. By the end of May 2020, there have been about 13K confirmed cases, and nearly 1K deaths.

SARS-CoV-2 is a member of Beta coronaviridae, single stranded positive sense RNA viruses which are enveloped viruses that possess extraordinarily large single-stranded RNA genomes ranging from 26 to 32 kilobases in length (*Zhu et al*., *2020*)

Growing evidence of the limitations of qRT-PCR prompts further consideration of the limitations of this diagnostic test. First, there are already over 7 different SARS-CoV-2 nucleic acid PCR tests (*Wang et al*., *2020*), yet many variation in sensitivity have been seen which might be extrinsic to the kit construction. Thus information on virus stability with special regards to its thermal stability and viral integrity of the SARS-CoV-2 in the environment at different temperature conditions is important for understanding virus transmission, stability, integrity and handling the spacemen’s properly for molecular diagnoses of the disease.

In this study, we reported the thermal stability of different SARS-CoV-2 isolates at different temperatures/ time combination points ranging from 20 °C to 70 °C for 2, 4, and 6 hours as measured by quantitative real time RT-PCR.

## Materials and Methods

Virus manipulation and RNA extraction were performed in biosafety level three cabinet (germfree biosafety cabinet, SEA-III, 316 ss).

### Samples

Oropharyngeal swabs (*n=5*) were taken from recently infected military patients with COVID-19 from Egypt during April 2020. The samples transferred in transportation media (sterile MEM with ampicillin100IU/ml, streptomycin 100μg/ml) at 4 °C in biosafety transporting device to main chemical laboratories at ALMAZA-Cairo.

### Thermal treatment

All samples were tested first for the presence of SARS-COV2 virus using virasure^®^ RT-PCR kit (SARS-CoV-2 Real Time PCR detection KIT high profile. Cat # VS-NCO212H), positive samples were subjected to limited dilutions and aliquoted (140μl) in nuclease free 0.2ml PCR tubes and subjected to temperature of 20 °C, 30 °C, 40 °C, 40 °C, 60 °C and 70 °C for 2, 4 and 6 h. using T professional 3000 thermal cycler (Biometra).

### RNA extraction

RNA from thermally treated and untreated samples were extracted using RNeasy Mini Kit (Qiagen cat # 52904) according to the manufacture instruction. Briefly, samples were lysed with 560 μl of buffer AVL at room temperature for 10 min, then 560 μl of absolute ethanol was added and the whole solution was then placed in the QIA amp Mini column provided with the kit. The spin columns were centrifuged at 8000 RPM/2min/4°C and washed with 500μl of washing solution AW1 then AW2. The RNA was eluted in 50μl of the AVE elution buffer and stored at −80°C till used.

### Real time RT-PCR

It was done using SARS-CoV-2 Real Time PCR detection KIT high profile. (Cat # VS-NCO212H) according to the manufacture instructions, briefly, the master mix was rehydrated with 15μL of rehydration Buffer and 5 μL of RNA of thermally treated and untreated samples were added (samples were run in duplicates), positive and negative controls (provided with the kit) were included in each test to judge the quality of amplification. Real time RT-PCR was done using Ariamx thermal cycler (Agilent, Germany) with the following parameters, reverse transcription step at 45oC/15 minute, followed by initial denaturing and enzyme activation step at 95oC/2 minute, then 45 cycled of denaturing at 95°C/10 sec and annealing/extension 60 °C/50 seconds with florescence collected at the end of this step.

Results presented as the mean ΔC_q_ values of the triplicated for each sample (mean C_q_ of the thermally treated sample – Mean C_q_ of the thermally untreated sample) and considered negative when there is no amplification (C_q_ ≥45) which means that there is no amplification and the virus completely inactivated (lost its integrity)

### Statistical analysis

The ΔC_q_ values were tested for significant at CI of 95% (Two way ANOVA) using IPM-SPP package V21. Data presented as mean ΔC_q_ values of the triplicated of 5 samples tested at each temperature/time combination points. Data were graphically presented using graph pad Prism V8.0.2

## Results

Samples used in the current study were collected from clinically morbid patients with the classical symptoms of SARS-CoV-2 including hyperthermia (>38.5°C) dyspnea, dry cough, and atypical pneumonia as confirmed by chest X rays. Oropharyngeal swabs were taken and confirmed positive for the presence of the virus using qRT-PCR (fig1 and 2). All samples gave positive C_q_ value ranging from (21.56-28.10) for orf1 gene and (23.10 −29.42) for the N gene.

**Fig 1.**
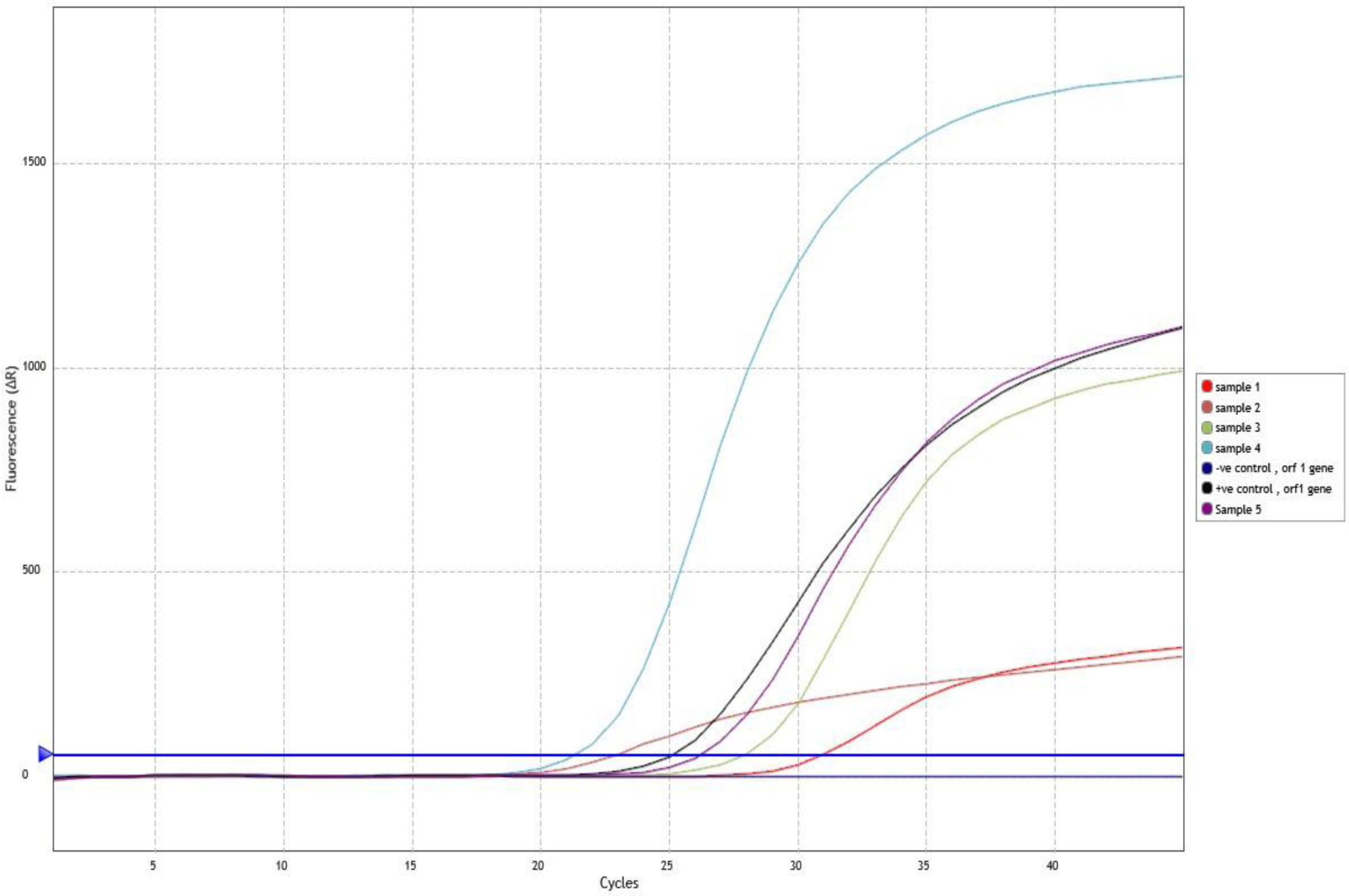
The amplification plot of samples taken from patient’s showed the amplification of the orf1 gene of SARS-CoV-2. Positive and negative controls were included in the reaction to judge the efficacy of the amplification.

**Fig 1.**
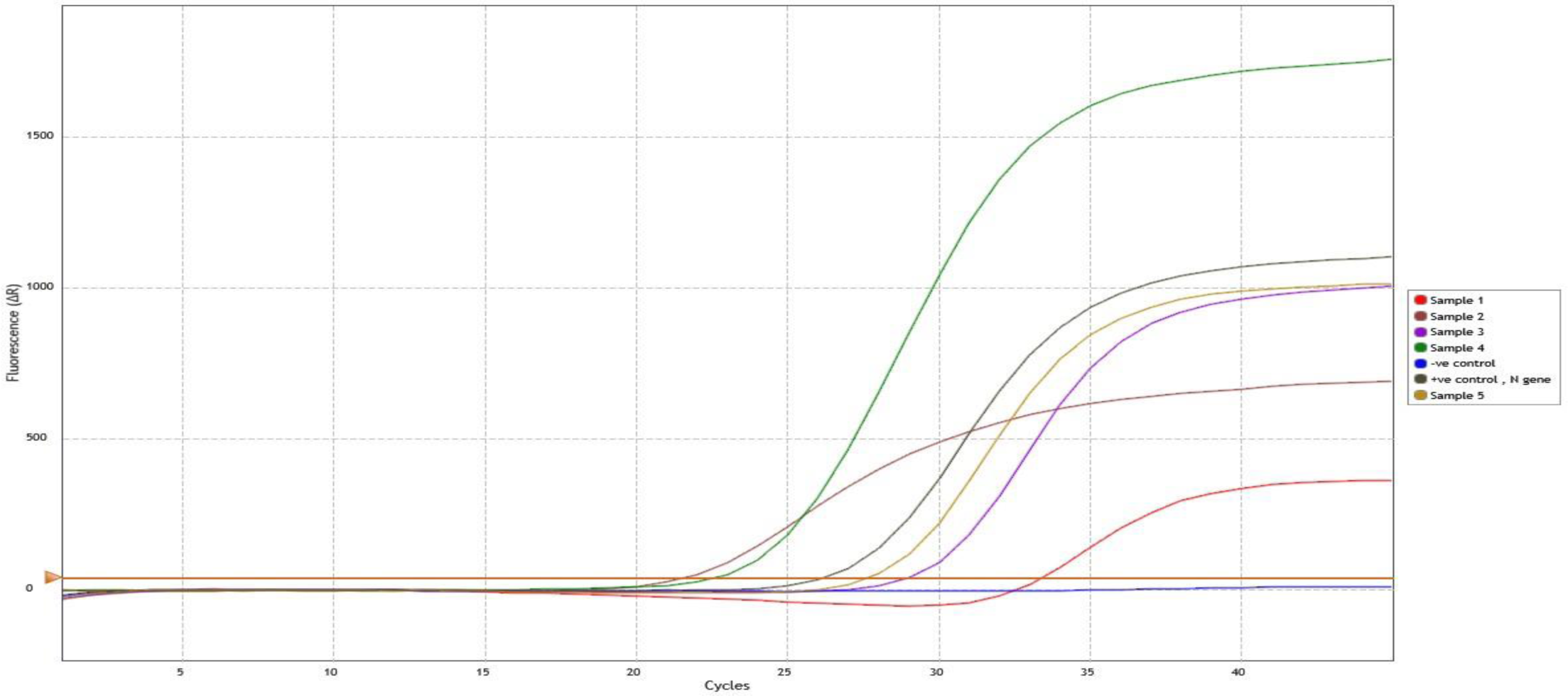
The amplification plot of N gene from morbid patients. Positive and negative controls were added at judge the amplification efficacy.

Samples with high viral titer were then aliquoted and thermally treated, the relative quantitation of both orf1 and N genes were done to examine the effect of each thermal/time combination point on the virus integrity. As then in table 1, fig (3 and 4) temp up to 40°C for up to 4 h did not significantly affect the mean relative C_q_ values for the orf1 gene were as at 40°C/6h the results became significantly (P ≤0.05) differed which means that the virus inactivation starts. At 50 °C the virus begin to lose its integrity. Complete virus inactivation was found at 60 °C/ 6h and at 70°C /2h.

**Fig (3).**
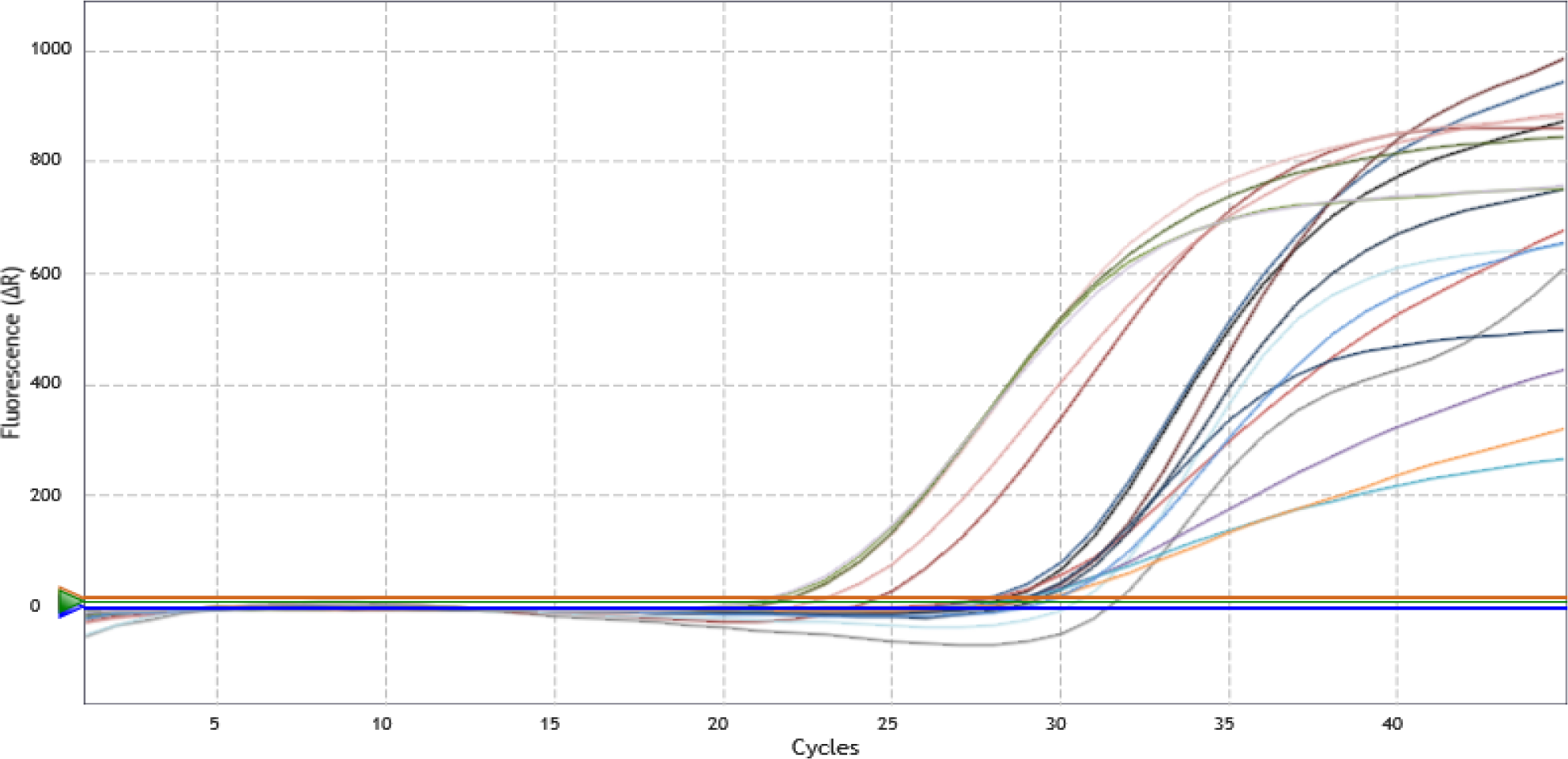
Representative amplification plot of orf1 and N genes for thermally treated samples at different thermal/time points. The ΔC_q_ values were presented in table 1.

**Fig (4).**
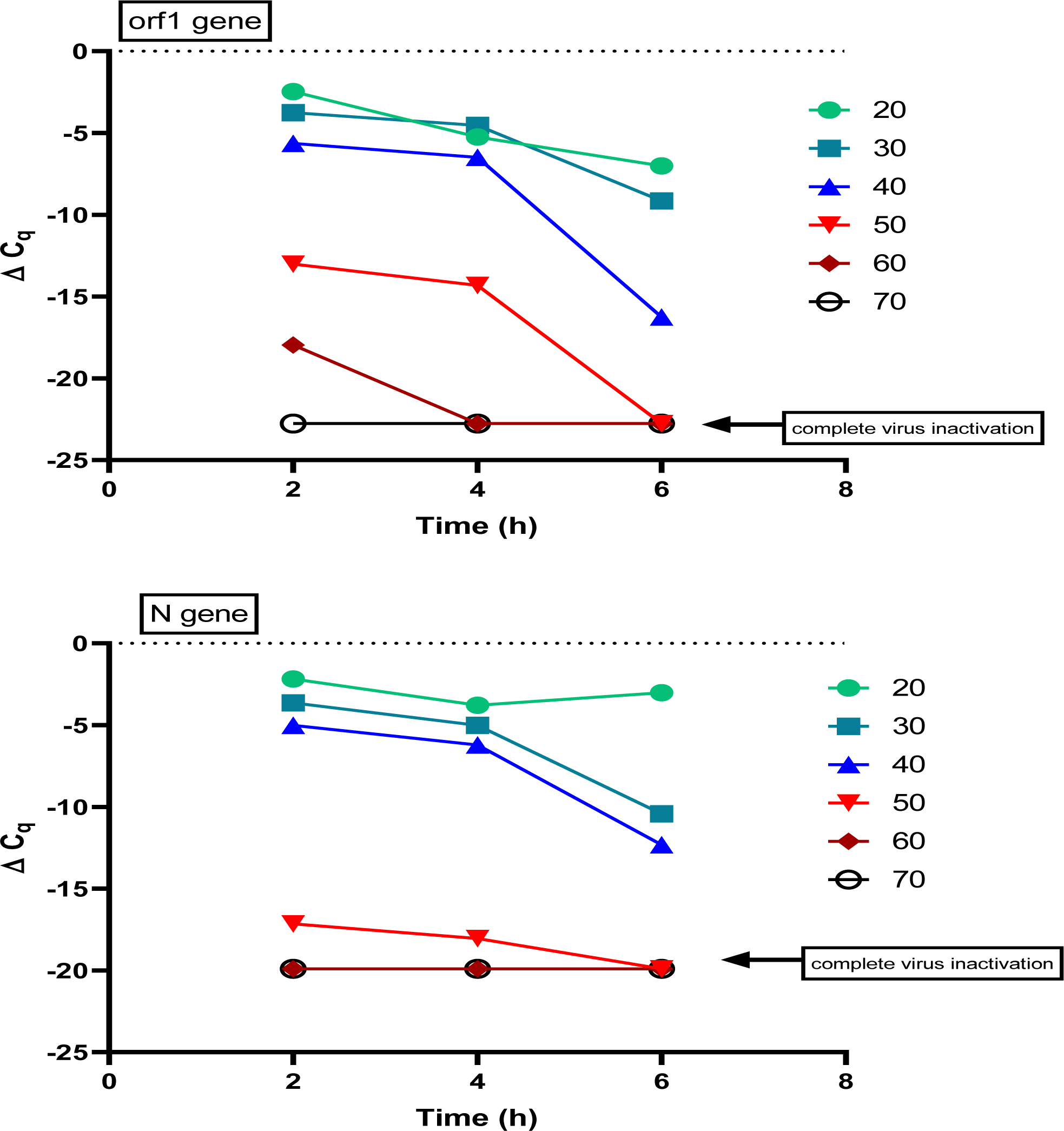
The effect of temp/time combination on the SARS-CoV-2 as measured by qRT-PCR for both orf1 and N genes. Data represented as the ΔC_q_ values (as shown in table 1).

**Table (1).** Stability of SARS-CoV-2 as measured by quantitative real time PCR Results expressed as mean ΔC_q_ value of the replicates ±SEM

| Treatment<br>temp | orf1 gene |  |  | N gene |  |  |
| --- | --- | --- | --- | --- | --- | --- |
|  | Treatment time ( h) |  |  |  |  |  |
|  | 2h | 4h | 6h | 2h | 4h | 6h |
| 20 | -2.47±0.1* | -5.24±0.02 | -7.01±0.5 | -2.18±0.1* | -3.79±0.02 | -3.02±0.5 |
| 30 | -3.75±0.4 | -4.54±0.03 | -9.15±0.3 | -3.64±0.4 | -5.05±0.03 | -10.43±0.3 |
| 40 | -5.64±0.3 | -6.49±0.04 | -16.26±0.4 | -5.02±0.3 | -6.21±0.04 | -12.32±0.4 |
| 50 | -13.01±0.3 | -14.32±0.05 | -22.76±0.2 | -17.15±0.3 | -18.05±0.05 | -19.9±0.2 |
| 60 | -17.96±0.2 | -21.96±0.2 | -22.76±0.2 | -19.9±0.2 | -19.9±0.2 | -19.9±0.2 |
| 70 | -22.76±0.2 | -22.76±0.2 | -22.76±0.2 | -19.9±0.2 | -19.9±0.2 | -19.9±0.2 |

\* All experimental studies were done in three independent triplicates.
† $\Delta C_q$ = (mean $C_q$ of the thermally treated sample – Mean $C_q$ of the thermally untreated sample)

For the N gene, nearly the same results obtained as the virus remain stable when subjected to 40 °C/4h, increase temp and /or time results in rapid deterioration of the virus integrity and sever decrease in the relative C_q_ value.

All tested 5 isolates showed no difference in thermal susceptibility as the variance in the slandered deviation were very low and there were no significant difference in the ΔC_q_ values

## Discussion

SARS-CoV-2 can remain infectious on inanimate surfaces at room temperature for up to 9 day (*Pan et al*., *2020*) At a temperature of 30 °C or more the duration of persistence is shorter (*Kampf et al*., *2020*). In the current study we discussed the possibility of thermal stability of SARS-CoV-2 in different temp as measured by qRT-PCR.

Virus inactivation as measured by real time PCR is a quick hazardless approach to test virus stability without the need for Tissue culture procedures. Thermal time combination treatment of the SARS-CoV-2 revealed that the inactivation of the virus begin at 40°C/6h, complete inactivation was achieved at 50°C/4h and 60°C/4h when measured by amplification of orf1 gene, whereas using N gene amplification, complete inactivation was achieved at 50°C/4 h and 60°C/2h. these results indicating that the SARS-CoV-2 as all other coronavirus are heat sensitive and can be activated at a temp of 50 °C. The same results were obtained when the infectivity of the virus was tested using TCID50 after incubation at different temperatures, (*Chin et al*., *2020*). The virus was highly stable at 4°C (there was only ∼0.7-log unit reduction of infectious titer on Day 14). With the incubation temperature being increased to 70°C, the time for virus inactivation was reduced to 5 minutes.(*Chin et al*., *2020*) whereas virus subjected to 56 °C /30 min resulted in complete loss of virus infectivity when measured by TCID50.

Studies on the infectious bronchitis virus (one of the avian viruses belonging to the gamma coronaviridae) revealed that the virus is highly thermal unstable at temp of 50 °C and it needs a high salt concentration (nearly 1M of Na_2_Hpo_4_) to maintain its infectivity, yet it became completely in-infective at 60 °C. (*Hopkins, 1967*). Other studies on avian coronaviridae showed difference in thermal stability between different strains (*Otsuki et al*., *1979*), although most strains were completely inactivated at 45 °C30 min some isolates like IB-41 and IB-KH maintained very low activity even at a temp beyond 60°C/1h.

In our study we could not find any thermal stability variations among the tested 5 isolates indicating that the SARS-CoV-2 may not yet evolve to maintain its infectivity at a higher temp outside the host cells.

## Conclusion

Our study revealed that thermal inactivation of SARS-CoV-2 can be achieved at 50°C/6h or 60 °C/2h and that, the detection of SARS-CoV-2 by RT-PCR does not affected by lower temperature / time combination. Variation in thermal stability did not recognized among the 5 tested isolates of SAES-CoV-2 from patients in Egypt. Further investigation using cell culture technique for measuring the infectivity of SARS-CoV-2 at the corresponding temp/time combinations might be required for in depth analysis.

## Data Availability

data availability with contact my mail

